# A cohort study of the effect of SARS-CoV-2 point of care rapid RT-PCR at the Emergency Department on targeted admission

**DOI:** 10.1101/2021.12.29.21268501

**Authors:** Susanne E Mortazavi, Malin Inghammar, Claus Christiansen, Anne-Katrine Pesola, Mikael Stenkilsson, Magnus Paulsson

**Affiliations:** Lund University, Skåne University Hospital, Department of Laboratory Medicine, Division of Clinical Chemistry and Pharmacology, Lund, Sweden; Lund University, Skåne University Hospital, Department of Clinical Sciences Lund, Section for Infection Medicine, Lund, Sweden; Clinical Microbiology, Laboratory medicine Skåne, Region Skåne; Department of Emergency and Internal Medicine, Skåne University Hospital, Lund, Sweden

**Keywords:** Infectious disease, Virology, Emergency medicine, Hospital infection, Microbiology, Medical error, patient safety, quality of care

## Abstract

(ii)

**Background:** To prevent nosocomial transmission of SARS-CoV-2, infection control measures are implemented for patients with symptoms compatible with COVID-19 until reliable test results are available. This delay targeted admission to the most appropriate ward based on the medical condition. SARS-CoV-2 rapid antigen detection (RAD) tests and point of care (POC) rapid RT-PCR were introduced at emergency departments (EDs) in late 2020, but the consequence on targeted admission is unknown.

**Objectives:** To assess the effect of RAD tests and POC rapid RT-PCR (VitaPCR, Credo Diagnostics, Singapore) on targeted admission.

**Methods:** Patients presenting at the ED of a referral hospital (N = 2,940) between 13-Nov-2020 and 12-Jan-2021 were included. The study period was delimited by introduction of RAD tests and VitaPCR. Participant data was collected retrospectively, and outcome variables were length-of-stay (LoS), intrahospital transfers and targeted admission to COVID-19 ward.

**Results:** RAD tests reduced ED LoS for participants with positive tests or that were not tested. Negative VitaPCR results reduced mean hospital LoS by 1.5 (95%CI: 0.3–2.7) days and admissions to COVID-19 wards from 34.5 (95%CI: 28.9-40.5) to 14.7 (95%CI: 11.1-19.1) per 100 admissions. Introduction of VitaPCR reduced transfers between hospital wards in the first 5 days from 50.0 (95%CI: 45.0-55.0) to 34.0 (95%CI: 30.3-37.9) per 100 admissions.

**Conclusion:** RAD tests enabled rapid detection of SARS-CoV-2 infection which had pronounced effects on LoS at the ED. VitaPCR added the possibility of exclusion of the infection which increased targeted admissions, reduced intrahospital transfers and lead to shorter stay at the hospital.

## Introduction

The coronavirus disease 2019 (COVID-19) pandemic is caused by the Severe Acute Respiratory Syndrome Coronavirus 2 (SARS-CoV-2) that emerged in China in late 2019 (1). According to WHO, on 7 November 2021 over 248,467,363 global cases and 5,027,183 global deaths have been verified (2). Rapid detection and isolation of infected individuals are important to limit the spread of the virus and to protect patients and health care workers (3). Real-time reverse transcription polymerase chain reaction (RT-PCR) is the gold standard for SARS-CoV-2 detection, due to high sensitivity and specificity compared to other diagnostic methods (4). However, RT-PCR is time consuming and requires specialized laboratory settings, personnel, and instruments. As a less expensive and faster point-of-care test method, SARS-CoV-2 rapid antigen detection (RAD) tests became widely available during the autumn of 2020. However, RAD tests are generally inferior to RT-PCR in terms of sensitivity and specificity, which is particularly important when testing asymptomatic patients with low pretest probability when the main objective is to rule out infection (4-6).

In late 2020, the optimized point-of-care (POC) RT-PCR VitaPCR SARS-CoV-2 Assay (Credo Diagnostics Biomedical, Singapore) was introduced and implemented at the Skåne University Hospital, Lund, Sweden. The assay utilizes a single tube for collection of the nasopharyngeal swab, cell lysis and nucleotide extraction. The total analysis time is about 20 min. Sample preparation does not require specialized laboratory setting and the reported sensitivity and specificity for SARS-CoV-2 is 99,3% and 94,7%, respectively, possibly lower in samples with low viral load (7, 8).

During 2020, substantial reorganizations were made at Emergency Departments (ED) and hospital wards globally to cope with the extraordinary requirements caused by the SARS-CoV-2 pandemic, including infection prevention and control (IPC) precautions to prevent secondary cases among patients and hospital staff. The study region was largely affected by the second wave of the pandemic with a considerable increase of COVID-19 cases from 1 Sept 2020 – 31 January 2021 (9). Due to the broad clinical manifestations of COVID-19, infection cannot be safely excluded based on clinical symptoms and signs only (10). Before introduction of RAD tests and VitaPCR, all patients with suspected COVID-19 infection were isolated at the ED or admitted to hospital wards dedicated to patients with positive or unknown COVID-19 status until RT-PCR results from the core hospital facility were available. The typical time from sampling to results ranged between 12 and 24 h. In the high prevalence setting during the second wave of the COVID-19 pandemic, a large proportion of the patients at the ED met the definition of suspected COVID-19 and were admitted to COVID-19 isolation wards instead of targeted admission to wards specialized on the true medical problem. If the COVID-19 RT-PCR test was negative, IPC precautions were discontinued, and the patient transferred to another hospital ward for continued treatment.

Although the sensitivity and specificity of the VitaPCR is superior to that of any RAD test, and time to result is substantially shorter for VitaPCR than for RT-PCR tests analyzed at the core hospital laboratory, the effects of these improvements on patient care are unknown. We hypothesized that introduction of the faster tests in the algorithm facilitated clinical decisions at the ED, limited IPC precautions to when necessary and improved targeted admission. This study evaluates the introduction of RAD tests and VitaPCR based on length-of-stay at the ED and hospital ward, intrahospital transfers the first five days and targeted admissions to COVID-19 ward during the peak of the second wave.

## Materials and methods

### Study setting and design

This retrospective observational study is based on data from patients presenting at the Emergency Department, Skåne University Hospital, Lund, Sweden between Nov 13, 2020, and Jan 12, 2021. The hospital is a regional referral center, but the ED primarily serves the population of Lund and near surroundings (population of about 300,000). The total annual ED visits in 2020 were 59,000 patients. The present study was divided into three distinct time periods separated by the dates for introduction of RAD tests and VitaPCR, respectively: Period 1 (Nov 13 to Dec 2), Period 2 (Dec 3 to Dec 22) and Period 3 (Dec 23 to Jan 12). The standard of care from the beginning of the pandemic and Period 1 of the present study was analysis of nasopharyngeal samples for SARS-CoV-2 with RT-PCR at a core hospital laboratory facility (Laboratory medicine Skåne, Region Skåne, Lund, Sweden). On Dec 3, 2020 (Period 2 of this study), POC rapid SARS-CoV-2 antigen detection test (ClinitestRT; Siemens Healthineers, Erlangen, Germany) was introduced together with an algorithm to select which analysis method that was to be used. On Dec 23, POC testing with VitaPCR was added to the algorithm. The algorithms are presented in Figure 1. The standard of care for suspect or confirmed patients with COVID-19 were unchanged during the study period and no changes were made concerning routines for hospital admissions. The average weekly COVID-19 incidence rate in Skåne county per 100,000 inhabitants was 355 in Period 1, 623 in Period 2 and 630 in Period 3.

**Figure 1.**
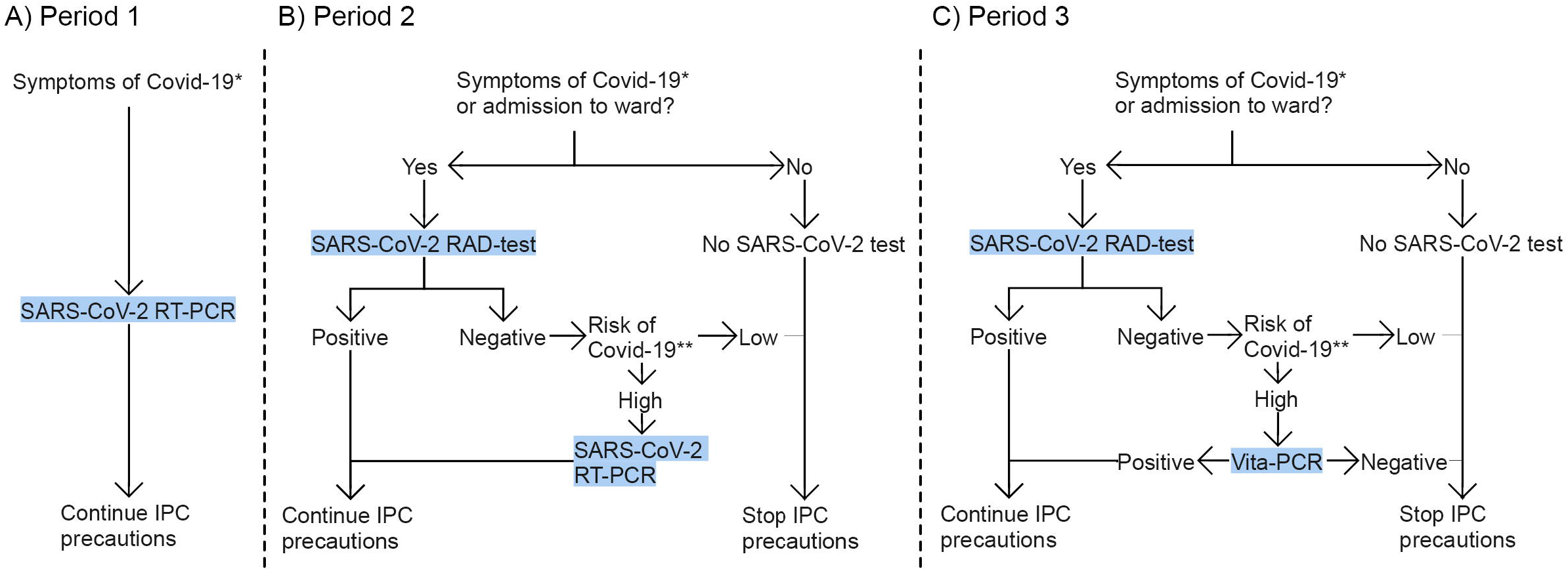
Algorithms for SARS-CoV-2 testing used at the Emergency Department during the study Period 1 (A), Period 2 (B) and Period 3 (C). During Period 1, patients with symptoms of COVID-19 were tested with RT-PCR and Infection prevention and control (IPC) precautions continued. During Period 2, a rapid antigen detection (RAD) test for SARS-CoV-2 was added, which enabled diagnosis of SARS-CoV-2 infection or discontinuation of IPC precautions for patients with low risk of SARS-CoV-2 infection and negative test result. During Period 3, the rapid POC RT-PCR Vita-PCR was introduced, and COVID-19 specific IPC precautions were ended for Vita-PCR negative patients regardless of symptoms. * newly developed upper or lower respiratory tract symptoms, fever, nausea, diarrhoea, malaise, anosmia or recent close contact with a person with confirmed SARS-CoV-2 infection. ** anosmia, fever with unknown origin, dyspnoea with unknown origin, cough or other respiratory symptoms and known contact with a COVID-19 case previous 7 days.

### Participants

All adult patients (≥ 18 years) that presented at the ED during the study period were screened for inclusion in the study. Inclusion criterions were any of the following: known COVID-19 infection at presentation to the ED, treatment in isolation room in the ED, emergency alerts labeled “Infection”, or emergency alerts with main complaints marked as “dyspnea”, “fever”, “infection”, “confusion”, “shock”, “cardiac arrest” and “non-specified illness”. All eligible patients during the study period were included.

### Variables

SARS-CoV-2 test results and test method were recorded and used for grouping of participants into independent variables: “Positive test at the ED” included participants that were sampled at the ED for a test that became positive for SARS-CoV-2 by RT-PCR, RAD test or VitaPCR, whereas “Negative test at the ED” included participants who tested negative at the ED, regardless of when the result of the test was given. Participants with positive SARS-CoV-2 analyzed before presentation to the ED were not sampled for SARS-CoV-2 and labeled “Positive test before admission”. Finally, “Not tested” included participants not tested for SARS-CoV-2 at the ED and the medical records did not specify COVID-19 status. The ICD-10 diagnoses at discharge from the ED and hospital wards were recorded and aggregated into compound variables (Supplemental file 1). Data on admission and transfer between wards were recorded and wards were grouped in the compound variables “COVID-19 ward” which were designated wards for patients with suspect or diagnosed COVID-19, “Mixed COVID-19/Internal Medicine ward” which were wards with ICP facilities but not dedicated only for patients with COVID-19, “Intensive care unit” (ICU) or “Other”, which included a broad range of specialized hospital wards. For each subject, sex and age were recorded and presented together with diagnoses as descriptive data. In all analyses, study period 1-3 were considered for exposure variables. Outcome variables were ED Length-of-Stay (LoS), discharge to home from ED, admission to hospital ward, hospital LoS, intrahospital transfers and targeted admissions.

### Data sources

Data on visits to the ED and hospital ward admissions were collected from the hospital records using key word search.

### Statistical analyses

Continuous variables with normal distribution are expressed as mean ± standard deviation (SD) or 95% confidence intervals (CI). Categorical variables are presented as counts and percentages. Comparisons were done by contingency tables with 95% confidence intervals (CI) reported, by One-Way-ANOVA with Tukey’s multiple comparison tests and by Fisher’s exact test. Analyses were performed with GraphPad Prism 9.0 (GraphPad Software, San Diego, CA, USA). P-values□<□0.05 were considered statistically significant. Missing data is presented in the tables.

### Ethical considerations

The study protocol has been reviewed and approved by the Swedish Ethical Review Authority (Dnr: 2021-00475). Informed consent was not retrieved according to Swedish legislation.

## Results

### Study cohort and participant enrollment

9,325 patients visited the Emergency Department during the study period, 2,940 of these met the inclusion criteria and were selected for enrollment in the study. There was a consecutive increase in the total number of patients that visited the ED and in the proportion that met the inclusion criteria: In Period 1: 781 participants out of 3,024 patient visits (25.8%) met the criteria, in Period 2: 988 participants out of 3,149 patient visits (31.4%), and Period 3: 1,171 participants out of 3,152 patient visits (37.2%). The mean age was 60.8 (SD ±20.8) years and 1497 (50.9%) of the participants were women.

### SARS-CoV-19 testing and test results

During the study period, a total of 1,866 (63.5%) participants were tested for COVID-19 at the ED. There was no significant difference in testing percentage between men and women (48.3% vs 51.7%). The mean age among participants that were tested for SARS-CoV-2 was 64.1 (SD ± 20.0) years and the mean age among those not tested was 55.1 (SD ± 21.0) years. As the study periods were defined by changes in testing routines, substantial differences in SARS-CoV-2 analysis methods were observed between the periods. Samples were analyzed with more than one method for 318 of the 2,940 participants (10.8%). The most common combination was RAD and RT-PCR (n = 186, 6.3%), followed by RAD and VitaPCR (n = 75, 2.6%), which is consistent with the testing algorithms in use during Period 2 respectively Period 3 for patients with negative RAD test but high risk of COVID-19 (Figure 1). After introduction of the RAD test and VitaPCR, there was a significant decrease in RT-PCR test analyses, but this method was still used in 9.5% (n = 111) of the 1,171 participants in Period 3 (Table 1).

**Table 1.** SARS-CoV-19 testing data. SARS-CoV-2 analysis methods used at the Emergency department. The use of Real-time polymerase chain reaction (RT-PCR) at the core laboratory decreased significantly between each study period as the point of care rapid antigen detection (RAD) test and point of care rapid RT-PCR VitaPCR were introduced in Period 2 and 3 respectively. Number of tests are presented with percentages (%) of total participants in each period and 95% confidence intervals (CI).

|  | **Period 1 (N = 781)** |  | **Period 2 (N = 988)** |  | **Period 3 (N = 1171)** |  | **Total (N = 2940)** |  |
| --- | --- | --- | --- | --- | --- | --- | --- | --- |
|  | n = | % (95% CI) | n = | % (95% CI) | n = | % (95% CI) | n = | % (95% CI) |
| **Analysis method** |  |  |  |  |  |  |  |  |
| RT-PCR | 443 | 56.7 (53.2 - 60.2) | 298 | 30.2 (27.4 - 33.1) | 111 | 9.5 (7.9 - 11.3) | 852 | 29.0 (27.4 - 30.6) |
| RAD test | 1 | 0.1 (0.007 - 0.7) | 506 | 51.2 (48.1 - 54.3) | 395 | 33.7 (31.1 - 36.5) | 902 | 30.7 (29.0 - 32.4) |
| VitaPCR | 0 | 0.0 (0.0 - 0.5) | 4 | 0.4 (0.2 - 1.0) | 435 | 37.1 (34.4 - 40.0) | 439 | 14.9 (13.7 - 16.3) |
| Total | 444 | 56.9 (53.4 - 60.3) | 650 | 65.8 (62.8 - 68.7) | 772 | 65.9 (63.2 - 68.6) | 1866 | 63.5 (61.7 - 65.2) |
| **Participants tested with multiple methods** |  |  |  |  |  |  |  |  |
| Antigen and RT-PCR | 0 | 0.0 (0.0 - 0.5) | 152 | 15.4 (13.3 - 17.8) | 34 | 2.9 (2.1 - 4.0) | 186 | 6.3 (5.5 - 7.3) |
| Antigen and Vita-PCR | 0 | 0.0 (0.0 - 0.5) | 0 | 0.0 (0.0 - 0.4) | 75 | 6.4 (5.1 - 8.0) | 75 | 2.6 (2.0 - 3.2) |
| VitaPCR and RT-PCR | 0 | 0.0 (0.0 - 0.5) | 2 | 0.2 (0.04 - 0.7) | 44 | 3.8 (2.8 - 5.0) | 46 | 1.6 (1.2 - 2.1) |
| RAD, VitaPCR and RT-PCR | 0 | 0.0 (0.0 - 0.5) | 2 | 0.2 (0.04 - 0.7) | 9 | 0.8 (0.4 - 1.5) | 11 | 0.4 (0.2 - 0.7) |

Of the 2,193 tests that were analyzed, 449 were positive (20.5%). The proportion of positive tests increased significantly during the study period (Period 1: 70 positive of 443 tested, 15.8%, 95% CI 12.7 – 19.4%; Period 2: 156 positive of 808 tested, 19.3%, 95% CI 16.7 – 22.2%; Period 3: 222 positive of 941 tested, 23.6%, 95% CI 21.0 – 26.4%). Of the 2,940 participants, 408 (13.9%) tested positive for SARS-CoV-2 infection at the ED, 1,458 (49.6%) tested negative, 568 (19%) had had a positive test before admission to the ED, and 506 (17.2%) were not tested for SARS-CoV-2 infection at the ED (Table 2).

**Table 2.**
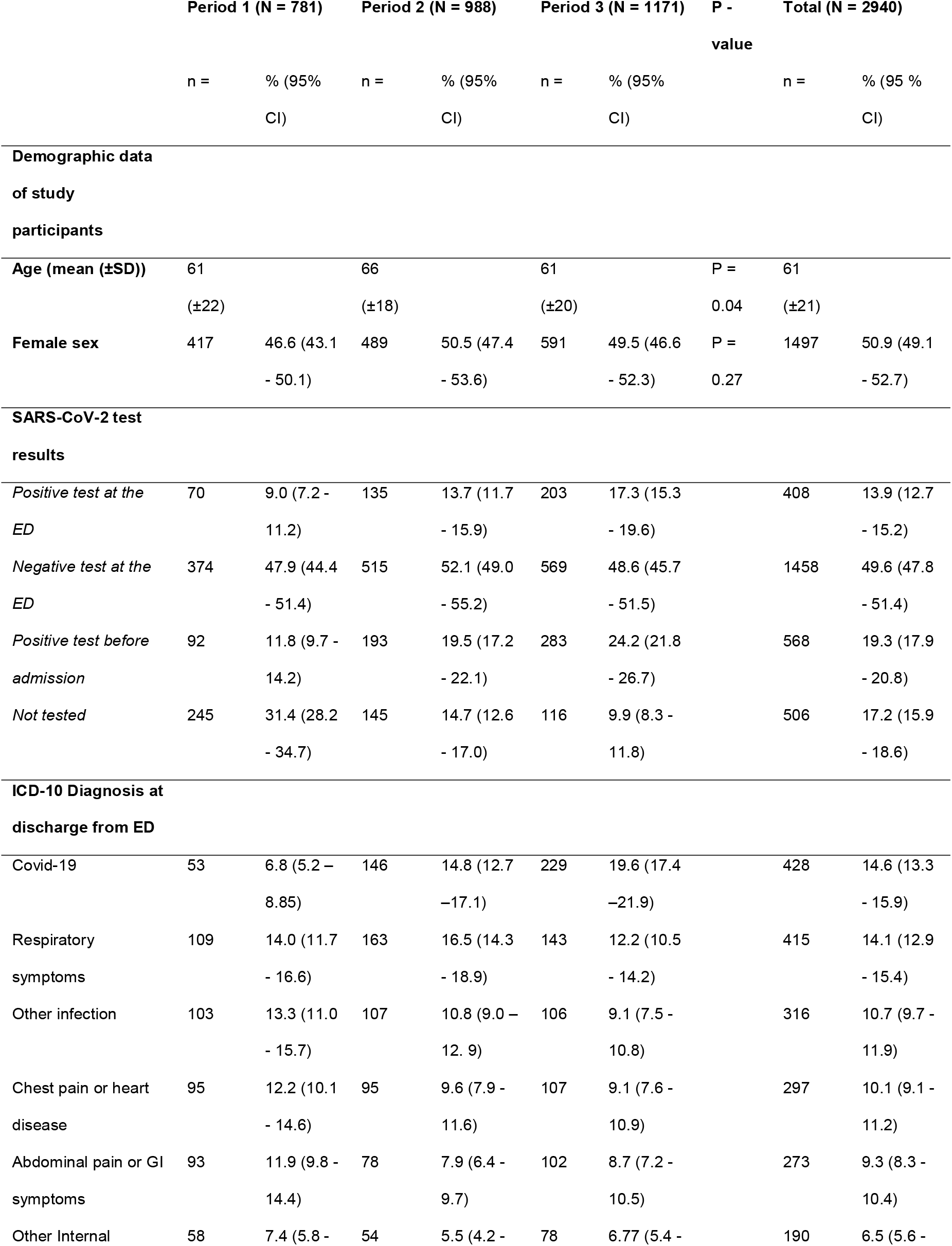
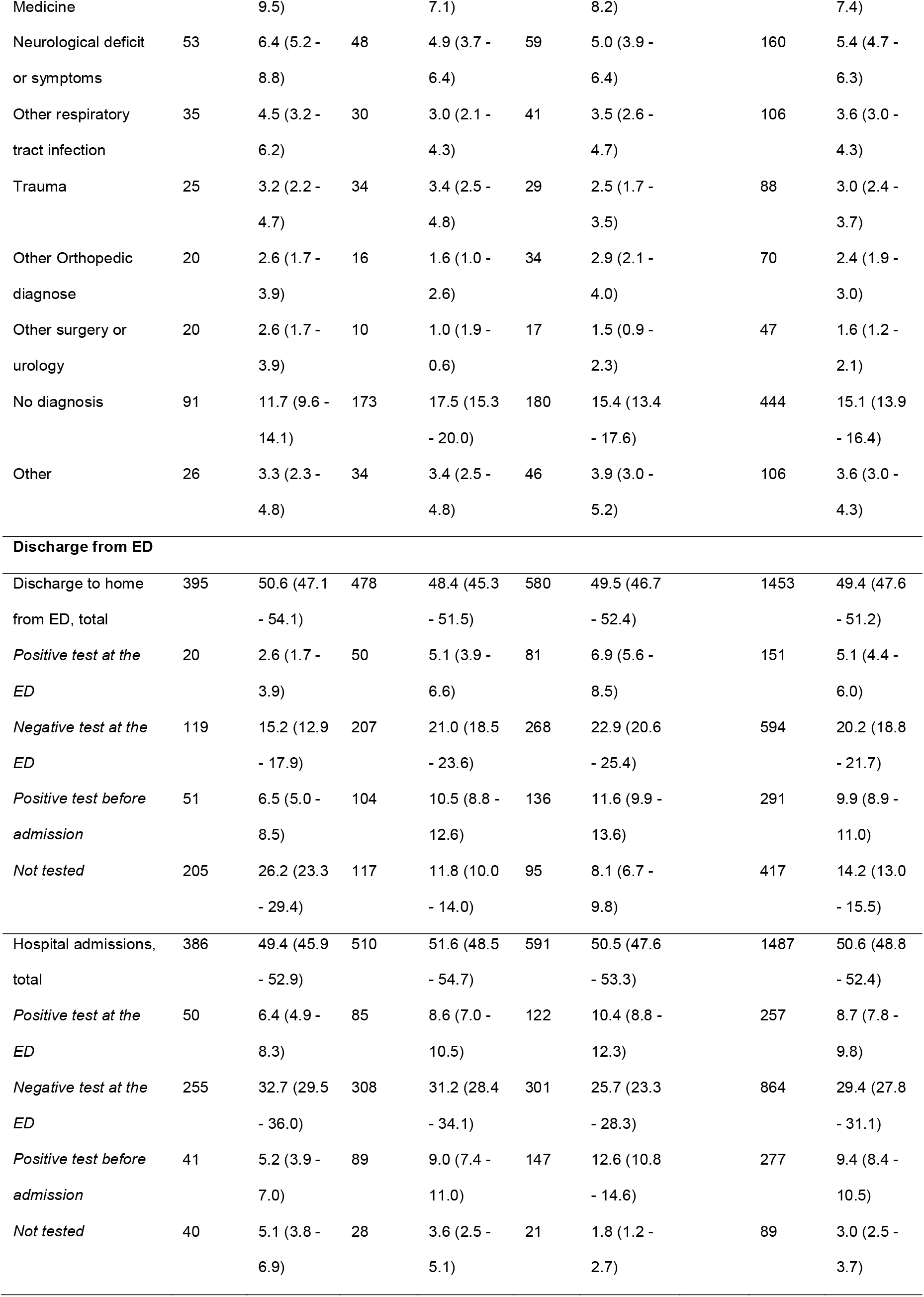
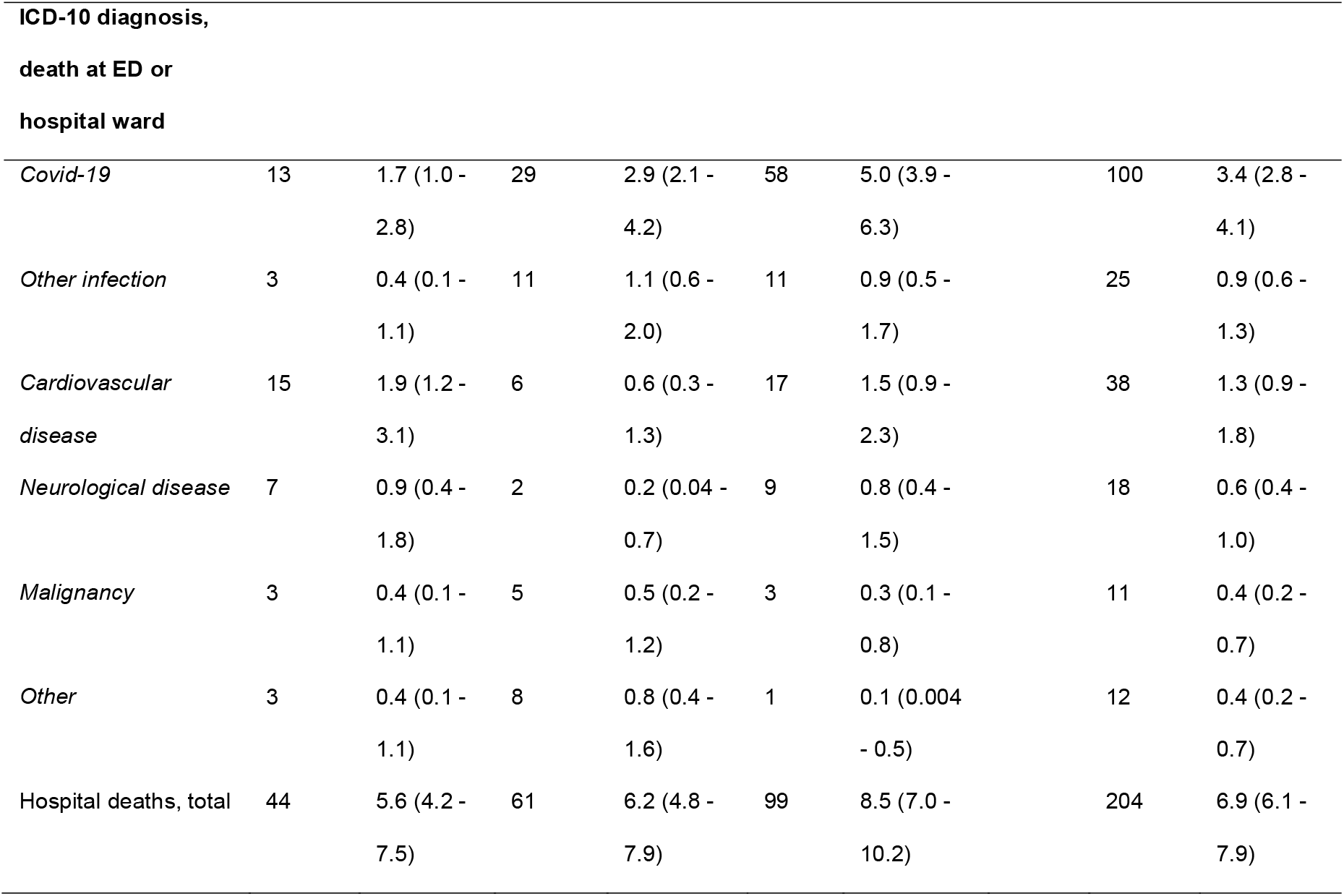
Participant characteristics at the ED. Characteristics of all study participants at the Emergency Department (ED). The percentage of participants with positive SARS-CoV-2 tests taken at the ED or before admission to the ED increased during the study period. ICD-10 diagnoses were grouped into compound variables (described in detail in Supplemental figure 1). COVID-19 diagnose increased, but no changes could be seen for other diagnoses. Total number of participants in each period (N=) and in each subgroup (n=) are presented with percentage of N= and 95% confidence interval (CI), or mean value with standard deviation (SD), as indicated. P-values calculated by One-Way-ANOVA with Tukey’s multiple comparison tests.

### Emergency Department Length-of-Stay

The mean Length-of-Stay (LoS) at the ED was 374 (SD ± 269) minutes (Table 3). This did not change significantly between the periods, but we observed a significant reduction in ED LoS between the three periods for participants with “Positive test at the ED” (P = 0.0002) or “Not tested” participants (P < 0.0001). Mean ED LoS for participants with “Positive test at the ED” decreased with 28 minutes (95% CI: 3.0 - 53.0) (P = 0.02) after introduction of RAD test, and another 15 min (95% CI: -7.6 - 37.6) after introduction of the VitaPCR. Participants that were “Not tested” for SARS-CoV-2 at the ED had a LoS at the ED that was reduced by 102 min (95% CI: 76.3 - 127.7) after introduction of the RAD test (P < 0.0001), which further decreased slightly (10 min; (95% CI: -33.16 - 13.16)) in Period 3.

**Table 3.**
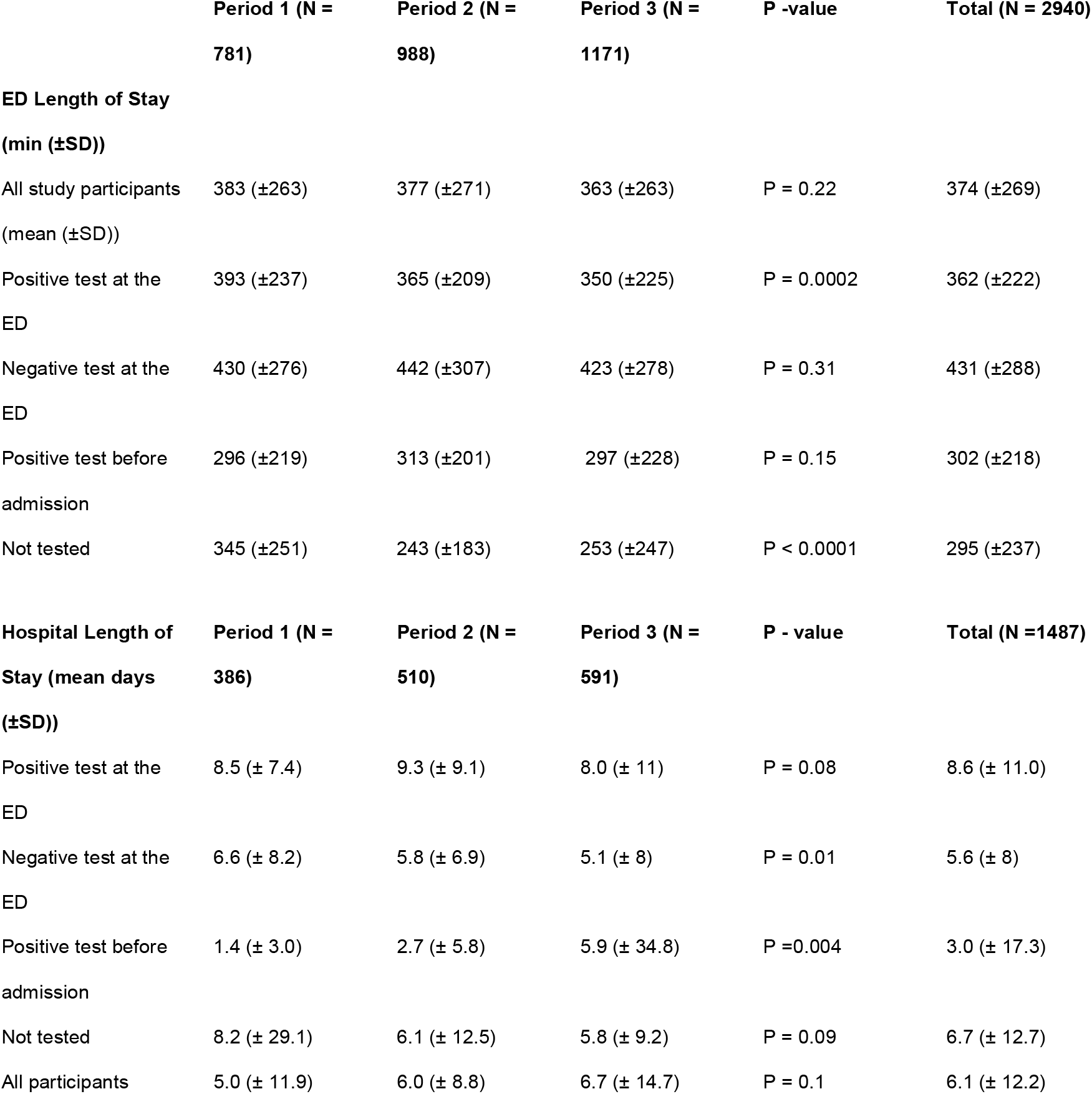
Length of Stay. Length of stay (LoS) in each study period at the Emergency department (ED) and total hospital stay. Numbers indicate mean minutes and standard deviation (SD) for the ED and mean days and SD for hospital LoS. P-value calculated by Fisher’s exact test.

### Emergency department discharge to home and hospital wards

COVID-19 diagnoses at discharge from the ED increased consecutively during the three study periods, from 6.8% in Period 1 (95% CI 5.2% - 8.85%), to 14.8% (95% CI: 12.7 -17.1) in Period 2 and 19.6% (95% CI: 17.4 -21.9) in Period 3. No statistically significant changes were seen for other diagnoses. Of the 2,940 participants, 1,487 (50.6%) were admitted to a hospital ward and 1,453 (49.4%) discharged to home. The percentage of participants that were discharged to home did not change notably during the study period. However, after introduction of RAD tests and VitaPCR, there was an increase of patients discharged to home from the ED for participants with either positive or negative test result at the ED or with a positive test before admission to the ED (Table 2). In Period 3, 47.1 % of participants with a “Negative test at the ED” were discharged to home, an increase from 31.8 % in Period 1 (P < 0.001) and 40.2 % in Period 2 (P = 0.02). Similarly, 37.0% of participants with a “Positive test at the ED” in Period 2 and 39.9% in Period 3 were discharged to home, compared to 28.6% in Period 1 (P = 0.65 and P = 0.11, respectively). Meanwhile, the percentage of hospital ward admissions of participants with “Negative test at the ED” was significantly reduced after introduction of the VitaPCR (Period 1: 32.7% (95 % CI: 29.5% - 36.0%); Period 2: 31.2% (CI 95% 28.4% - 34.1%); Period 3: 25.7% (CI 95% 23.3% - 28.3%). Conversely, the percentage of participants with “Positive test at the ED” of all hospital admissions increased from 6.4% (95% CI 4.9% - 8.3%) in Period 1, to 8.6% (95% CI 7.0% - 10.5%) in Period 2, to 10.4% (95% 8.8% - 12.3%) in Period 3.

### Hospital admissions and Length-of-Stay

We hypothesized that the total LoS for patients admitted to wards would be shortened by introduction of faster and more accurate testing methods that could safely exclude SARS-CoV-2 infection at the ED. The mean LoS during the entire study period was 6.1 (SD ± 11.0) days (Table 3). This increased slightly during the study period from 5.0 days in Period 1, 6.0 days in Period 2 to 6.7 days in Period 3 (P = 0.11). This increase was pronounced among participants that had a “Positive test before admission” to the ED for whom the mean hospital LoS increased from 1.4 (SD ± 3.0) days in Period 1 to 2.7 (SD ± 5.8) days in Period 2 and 5.9 (SD ± 34.8) (P < 0.004) days in Period 3. In contrast, there was a significant reduction in mean hospital LoS between the three periods for participants who had a “Negative test at the ED”. Introduction of the RAD test coincided with a reduction from 6.6 to 5.8 days (P = 0.27), and introduction of VitaPCR coincided with a further reduction to 5.1 days (Period 1 vs Period 3: P = 0.008; Period 2 vs Period 3: P = 0.046).

### Targeted admission and intrahospital transfers

The local routine was to discharge patients with suspect or confirmed COVID-19 from the ED to wards with facilities for IPC until test results. Patients with symptoms compatible with COVID-19 were transferred to other wards if the RT-PCR test was negative. However, negative RAD tests were not sufficient for termination of IPC measures in cases with high suspicion of COVID-19 as the accuracy of the test was unsatisfactory. While the trend for total hospital LoS was an increase, SARS-CoV-2 negative patients had a significant 1.5-day reduction in LoS after introduction of both tests. Hence, we hypothesized that introduction of the VitaPCR would increase targeted admission of participants with “Negative test at the ED”.

The proportion of participants that were admitted to COVID-19 wards, Mixed COVID-19/Internal medicine wards, ICU or Other wards did not vary significantly during the study period (Table 4). However, introduction of the algorithms including RAD test and VitaPCR were associated with a significant 32.0% decline in intrahospital transfers the first 5 days after admission (P < 0.0001). The number of participants with a “Negative test at the ED” that were transferred to another ward during the first 5 days of hospital admission decreased from 128 participants out of 386 (33.2%; 95% CI: 28.7% - 38.0%)) in Period 1, 122 participants out of 510 (23.9% (95% CI: 20.4% - 27.8%)) in Period 2 to 94 participants out of 591 (15.9% (95% CI: 13.2% - 19.1%)) in Period 3, a 52.1% reduction (Figure 2). There were no significant changes in intrahospital transfers between the periods for participants with positive tests or participants that were not tested.

**Table 4.** Descriptive and outcome data at hospital wards. The proportion of participants that were admitted different types of wards remained stable throughout the study period. Intrahospital transfers between wards the first 5 days after admission from the Emergency Department (ED) decreased after introduction of SARS-CoV-2 rapid antigen detection test (Period 2) and further with the introduction of the point of care rapid RT-PCR VitaPCR (Period 3). Hospital length of stay from admission at the ED to discharge to home from the hospital ward increased, particularly for patients with positive test before admission to the ED. Conversely, negative test at the ED coincided with a shorter length of stay in the latter two periods. Total number of participants in each period (N=) and in each subgroup (n=) are presented with percentage of N= and 95% confidence interval (CI), or mean value with standard deviation (SD), as indicated. P-values calculated by One-Way-ANOVA with Tukey’s multiple comparison tests.

|  | Period 1 (N = 386) |  | Period 2 (N = 510) |  | Period 3 (N = 591) |  | Total (N =1487) |  |
| --- | --- | --- | --- | --- | --- | --- | --- | --- |
|  | n = | % (95% CI) | n = | % (95% CI) | n = | % (95% CI) | n = | % (95 % CI) |
| **Admissions to ward** |  |  |  |  |  |  |  |  |
| *Covid-19 ward* | 166 | 43 (38.2 - 48) | 229 | 44.9 (40.6 - 49.2) | 274 | 46.4 (42.4 - 50.4) | 669 | 45 (42.5 - 47.5) |
| *Mixed Covid-19/Internal Medicine ward* | 86 | 22.3 (18.4 - 26.7) | 91 | 17.8 (14.8 - 21.4) | 98 | 16.6 (13.8 - 19.8) | 275 | 18.5 (29.8 - 34.6) |
| *ICU* | 10 | 2.6 (1.4 - 4.7) | 7 | 1.4 (0.7 - 2.8) | 6 | 1 (0.5 - 2.2) | 23 | 1.5 (1 - 2.3) |
| *Other* | 115 | 29.8 (25.4 - 34.5) | 166 | 32.5 (28.6 - 36.7) | 197 | 33.3 (29.7 - 37.2) | 478 | 32.1 (29.8 - 34.6) |
| *Missing data* | 9 | 2.3 (1.2 - 4.4) | 17 | 3.3 (2.1 - 5.3) | 16 | 2.7 (1.7 - 4.4) | 42 | 2.8 (2.1 - 3.8) |
| **Total** | **386** | **100.0 (99.0 - 100.0)** | **510** | **100.0 (99.3 - 100.0)** | **591** | **100.0 (99.4 - 100.0)** | **1487** | **100.0 (99.7 - 100.0)** |
| **Intrahospital transfers first 5 days** |  |  |  |  |  |  |  |  |
| *Positive test at the ED* | 35 | 9.1 (6.6 - 12.3) | 45 | 8.8 (6.7 - 11.6) | 55 | 9.3 (7.2 - 11.9) | 135 | 9.1 (7.7 - 10.6) |
| *Negative test at the ED* | 128 | 33.2 (28.7 - 38.0) | 122 | 23.9 (20.4 - 27.8) | 94 | 15.9 (13.2 - 19.1) | 344 | 23.1 (21.1 - 25.3) |
| *Positive test before admission* | 25 | 6.5 (4.4 - 9.4) | 27 | 5.3 (3.7 - 7.6) | 49 | 8.3 (6.3 - 10.8) | 101 | 6.8 (5.6 - 8.2) |
| *Not tested* | 3 | 0.8 (0.2 - 2.3) | 11 | 2.2 (1.2 - 3.8) | 3 | 0.5 (0.1 - 1.5) | 17 | 1.1 (0.7 - 1.8) |
| **Total** | **191** | **50.0 (45.0 - 55.0)** | **205** | **40.2 (36.0 - 44.5)** | **201** | **34.0 (30.3 - 37.9)** | **597** | **40.1 (37.7 - 42.7)** |
| **ICD-10 Diagnosis on discharge** |  |  |  |  |  |  |  |  |
| *Covid-19* | 81 | 21.0 (17.2 - 25.3) | 161 | 31.5 (27.6 - 35.7) | 241 | 40.8 (37.0 - 44.9) | 298 | 32.5 (30.1 - 34.9) |
| *Other infection* | 80 | 20.7 (25.0 - 25.0) | 80 | 15.7 (12.8 - 19.1) | 56 | 9.5 (7.4 - 12.1) | 208 | 14.5 (12.8 - 16.4) |
| Chest pain or heart disease | 43 | 11.1 (8.4 - 14.7) | 52 | 10.2 (7.8 - 13.1) | 48 | 8.1 (6.2 - 10.6) | 144 | 9.6 (8.2 - 11.2) |
| Other respiratory tract infection | 38 | 9.8 (7.3 - 13.2) | 50 | 9.8 (7.5 - 12.7) | 49 | 8.3 (6.3 - 10.8) | 136 | 9.2 (7.8 - 10.8) |
| Surgery or Urology | 33 | 8.5 (6.2 - 11.8) | 31 | 6.1 (4.3 - 8.5) | 48 | 8.1 (6.2 - 10.6) | 102 | 7.5 (6.3 - 9.0) |
| Other Internal Medicine | 31 | 8.0 (5.7 - 11.2) | 29 | 5.7 (4.0 - 8.0) | 38 | 6.4 (4.7 - 8.7) | 93 | 6.6 (5.4 - 8.0) |
| Neurological deficit or symptoms | 25 | 6.5 (4.4 - 9.4) | 28 | 5.5 (3.8 - 7.8) | 33 | 5.6 (4.0 - 7.8) | 75 | 5.8 (4.7 - 7.1) |
| Respiratory symptoms | 15 | 3.9 (2.4 - 6.3) | 23 | 4.5 (3.0 - 6.7) | 22 | 3.7 (2.5 - 5.6) | 52 | 4.0 (3.1 - 5.2) |
| Malignancy | 15 | 3.9 (2.4 - 6.3) | 12 | 2.3 (1.3 - 4.1) | 14 | 2.4 (1.4 - 3.9) | 38 | 2.8 (2.0 - 3.7) |
| Trauma | 7 | 1.8 (0.9 - 3.7) | 13 | 2.5 (1.5 - 4.3) | 11 | 1.9 (1.0 - 3.3) | 26 | 2.1 (1.5 - 2.9) |
| Abdominal pain and GI symptoms | 6 | 1.6 (0.7 - 3.3) | 9 | 1.8 (0.9 - 3.3) | 6 | 1.0 (0.5 - 2.2) | 17 | 1.4 (0.9 - 2.1) |
| Other Orthopedic diagnose | 0 | 0.0 (1 - 0.0) | 1 | 0.2 (0.01 - 1.1) | 2 | 0.3 (0.1 - 1.2) | 13 | 0.2 (0.1 - 0.6) |
| No diagnosis | 9 | 2.3 (1.2 - 4.4) | 11 | 2.2 (1.2 - 3.8) | 12 | 2.0 (1.2 - 3.5) | 30 | 2.2 (1.5 - 3.0) |
| Other | 3 | 0.8 (0.2 - 2.3) | 11 | 2.2 (1.2 - 3.8) | 10 | 1.7 (0.9 - 3.1) | 14 | 1.6 (1.1 - 2.4) |

**Figure 2.**
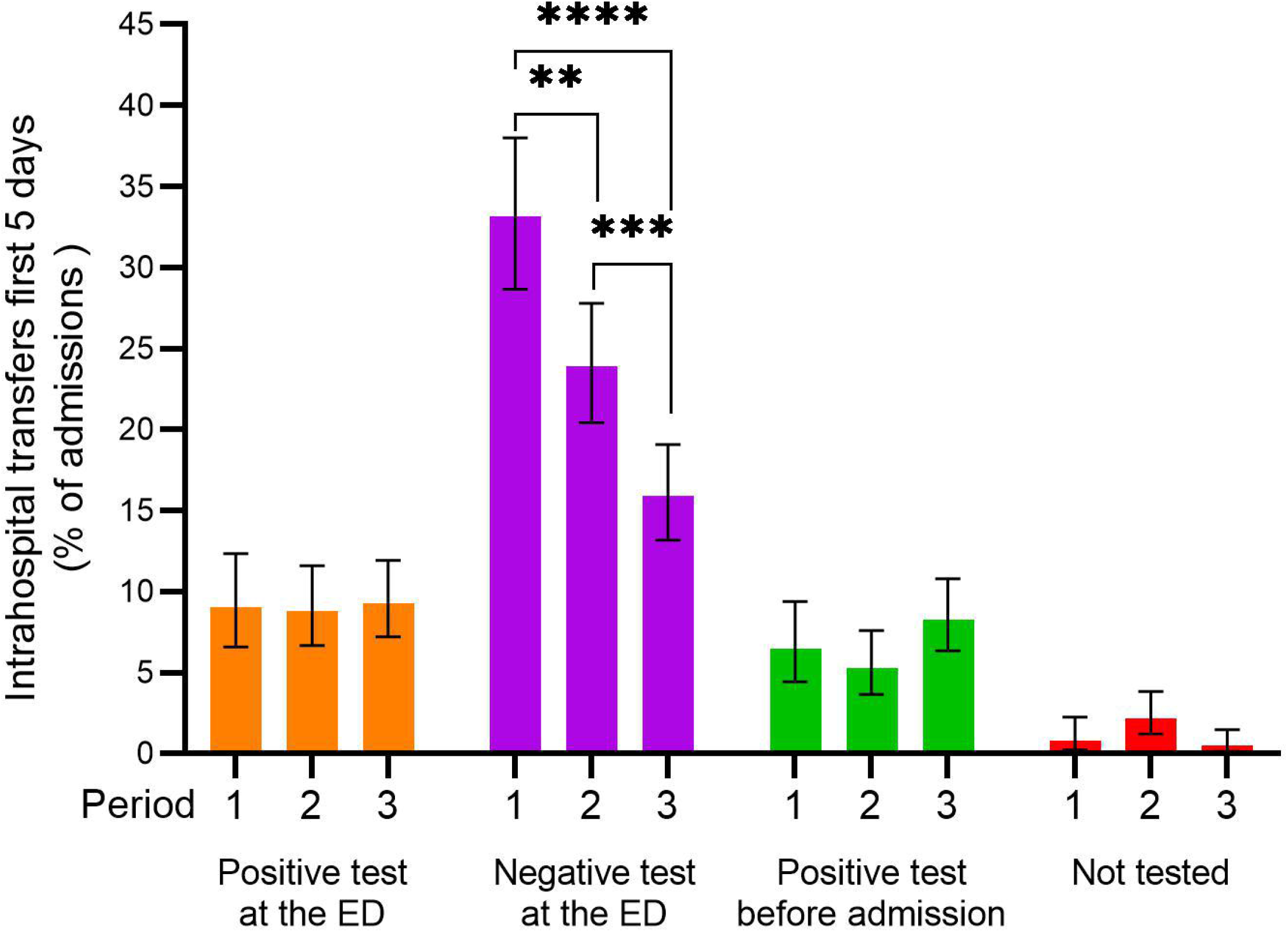
Intrahospital transfers first 5 days after hospital admission. Percentage of admitted participants that were transferred between hospital wards during the first 5 days of admission from the emergency department (ED). Participants that had a negative SARS-CoV-2 test at the ED were less likely to be transferred in Period 2 and 3, after introduction of the SARS-CoV-2 rapid antigen detection test and VitaPCR respectively. Bar shows mean value in percent and bars 95 % confidence interval. ** indicate P ≤ 0.01, *** P ≤ 0.001 and **** P ≤ 0.0001, calculated with Fisher’s exact test.

Similarly, participants with “Negative test at the ED”, were 57% less likely to be admitted to a COVID-19 ward and 81% more likely to be admitted to a targeted hospital ward after the introduction of both the RAD test and VitaPCR (Table 5). For participants with positive tests before or at the ED, or that were not tested, no significant changes were seen during the study period.

**Table 5.**
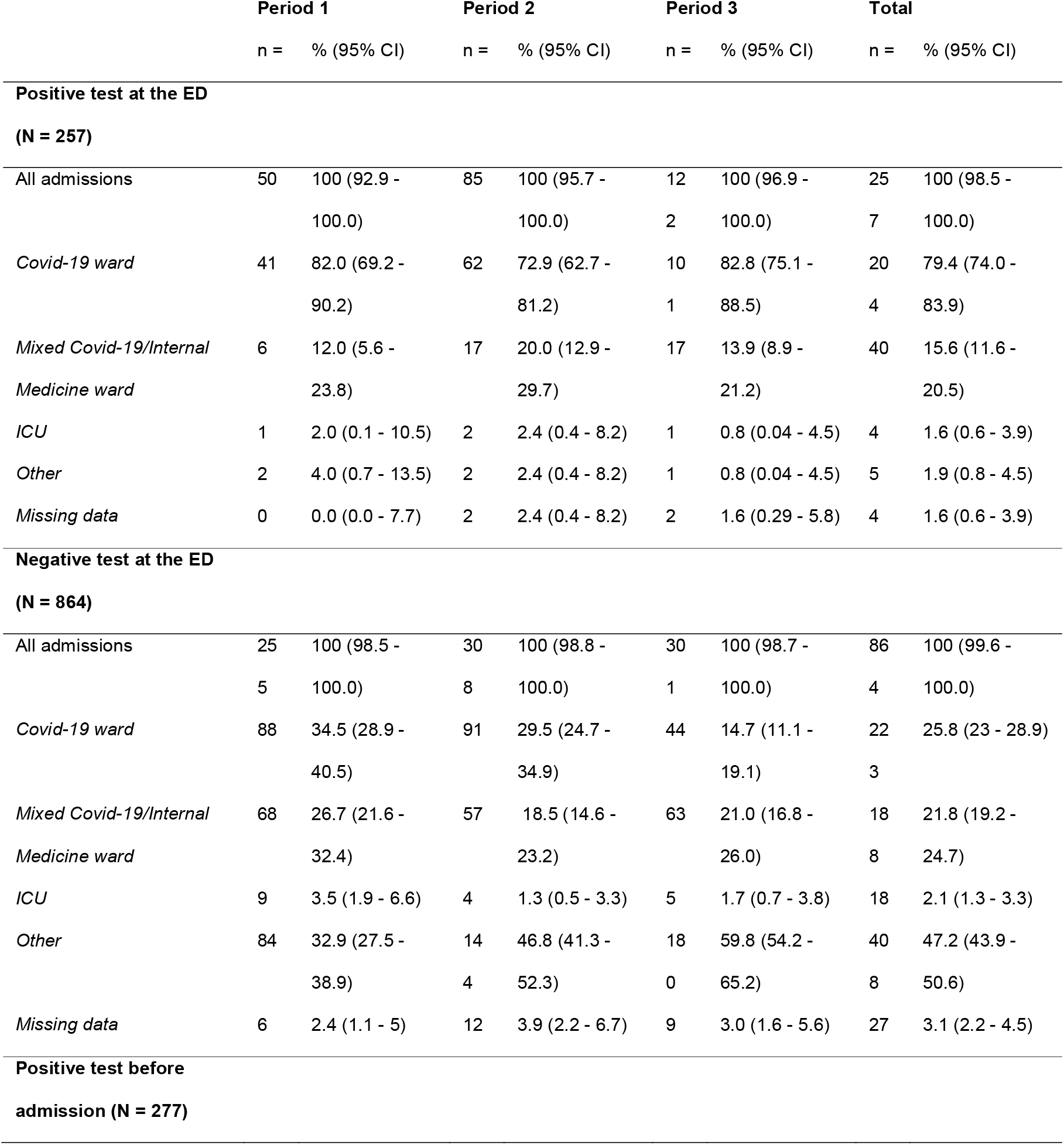
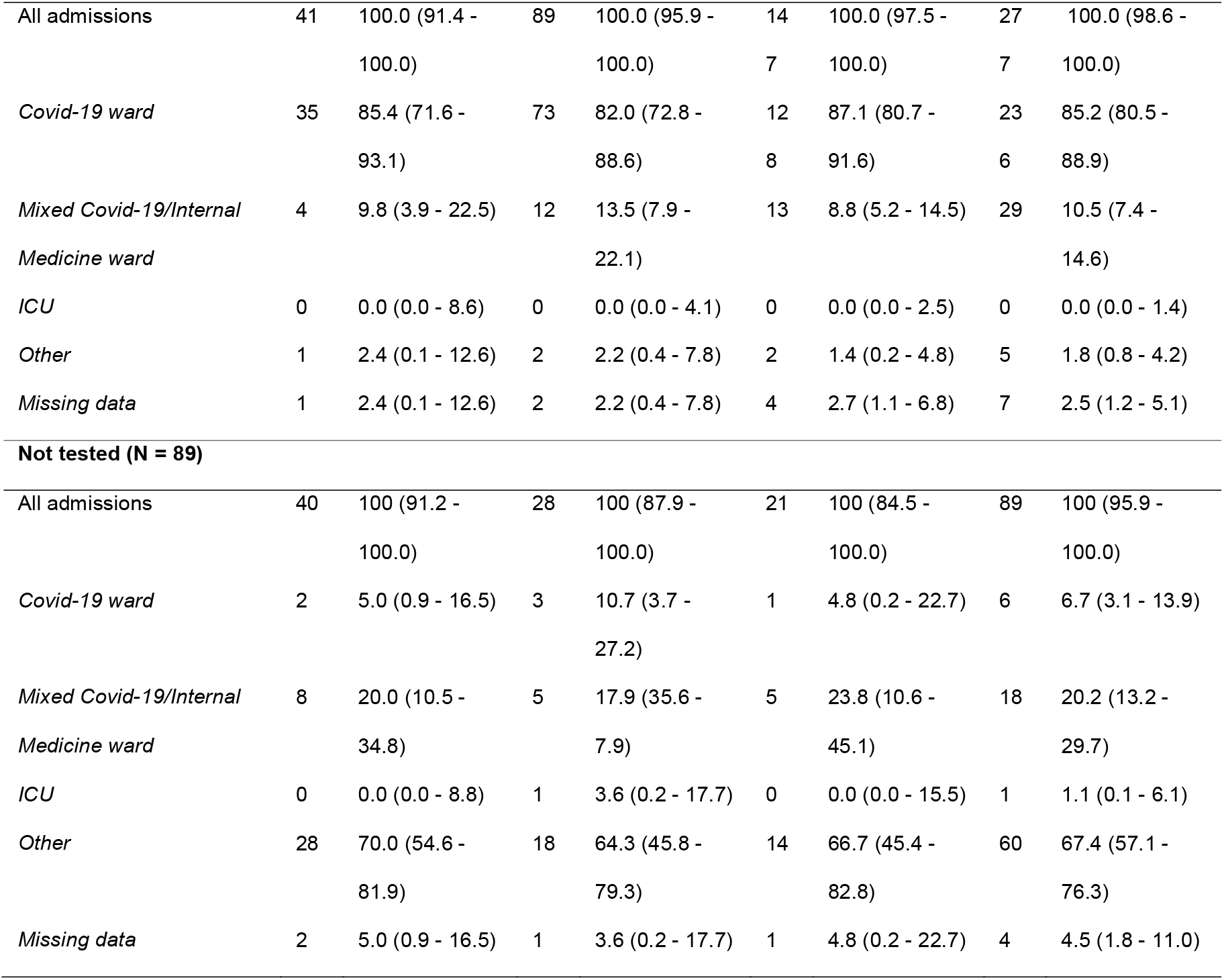
Targeted admissions. Targeted admission of patients with positive test or suspect SARS-CoV-2 infection to a COVID-19 ward or other ward with infection prevention control (IPC) facilities did not change by the introduction of rapid point of care tests (Period 2: Rapid antigen detection test; Period 3: rapid RT-PCR VitaPCR) at the Emergency Department (ED). However, test-negative patients were increasingly admitted to appropriate specialized hospital wards (“Other”) during Period 2 and 3. Total number of participants in each period (N=) and in each subgroup (n=) are presented with percentage of N= and 95% confidence interval (CI).

## Discussion

In this retrospective study, we have explored the impact of the introduction of SARS-CoV-2 RAD tests and the POC rapid RT-PCR VitaPCR on patient care during a period of high prevalence of SARS-CoV-2 infection. The study included 2,940 participants that visited the ED, who were grouped into three periods to highlight differences between the previous standard of care (Core laboratory RT-PCR) in Period 1, introduction of RAD test in Period 2 and introduction of VitaPCR in Period 3. Importantly, the results reveal that the implementation had a significant effect on length of stay at the ED and hospital, intrahospital transfers first 5 days and targeted admission to wards with IPC facilities, which has implications for the treatment of the individual patient, patient safety, hospital infection control and optimal resource use.

Before the introduction of point-of-care rapid SARS-CoV-2 tests at the ED, patients with suspected COVID-19 infection were admitted to COVID-19 diagnostic and treatment wards with IPC facilities until results from RT-PCR became available. Patients with negative tests were then transferred to the most appropriate ward based on the medical need. While this prevented secondary cases of COVID-19, intrahospital transfers have been related to unfavorable events such as increased falls, medication errors, length-of-stay and hospital-acquired infections (11, 12), as well as increased nurse and doctor work load (13). This study analyzed targeted admission to COVID-19 or non-COVID-19 wards directly, as well as using intrahospital transfers and hospital time of stay as surrogate markers of targeted admission. A significant improvement was seen in the outcome of all these variables that started with the introduction of the RAD test but was further pronounced after addition of the VitaPCR. Negative RAD test led to cessation of IPC at the ED and before admission to a hospital ward for patients without high risk of COVID-19. However, during Period 2, it was still necessary to exclude SARS-CoV-2 infection with RT-PCR at the core laboratory for patients with high risk of the infection. In contrast, in Period 3 a negative VitaPCR was sufficient for cessation of IPC even for patients with symptoms compatible with COVID-19. Hence targeted admission was possible for negative patients regardless of symptoms, leading to fewer intrahospital transfers, immediate initiation of appropriate therapy and shorter length-of-stay at the hospital. No significant effect on targeted admission variables could be seen for test positive participants, likely because both suspected and confirmed cases of COVID-19 were admitted to wards with IPC measures. In fact, hospital LoS increased substantially for patients with positive SARS-CoV-2 test before admission to the ED. The reason for this is unknown as no difference in age or in proportion that received ICU care (data n.s.) were seen between the periods.

High patient load at the ED puts increased pressure on hospital wards, which can lead to ED crowding and decreased patient safety (14). A key characteristic of crowding is an extended mean ED length-of-stay. The overall ED LoS did not substantially change during the study period, which is notable considering the increased number of patients in the latter periods of the study. Even though the proportion of COVID-19 diagnosis increased in the ED, participants with positive rapid tests at the ED (RAD test or VitaPCR) spent a shorter time at the ED than in Period 1. Similarly, it is interesting to note that discharges from the ED to home increased during the study period, particularly for participants that were tested for SARS-CoV-2 at the ED. The change was seen after introduction of the RAD tests, possibly because of the faster diagnostic work up that was enabled by the rapid tests. In the algorithms used during the study period, a positive RAD test was enough to confirm COVID-19. Hence, it can be expected that any change in patient care inflicted by the rapid tests for participants that tested positive at the ED would occur already in Period 2 (after introduction of RAD test) and not significantly changed after introduction of the VitaPCR.

The study period is unique in that allows for interrogation of the sequential introduction of two rapid POC analysis methods with direct implications for IPC management of patients in a high endemic setting. The study site is the only ED in the area and the hospital treats all kinds of medical emergencies, which resulted in an unbiased adult population that is generalizable to similar ED’s. The study population was sufficiently large for the proposed analysis. The conclusions drawn here may well be applicable in other similar settings, for instance the need for rapid and accurate diagnostic tools for the annual influenza virus epidemic, which put similar demands on ED and hospital wards. Limits of the present study mainly relate to the retrospective study design. At the time of introduction of these diagnostic methods, it was not possible, or ethically defendable, to conduct a prospective randomized study.

## Conclusion

In conclusion, the implementation of the testing algorithm including VitaPCR enabled exclusion of SARS-CoV-2 infection in patients at the ED, which reduced intrahospital transfers, shortened the stay at hospital wards, and increased targeted admissions to an appropriate ward. In addition, early identification of SARS-CoV-2 infection with rapid tests at the ED facilitated decisions at the ED and reduced ED time-of-stay. It would be of great interest to further investigate the health-economic implications of these results.

## Data Availability

All data produced in the present study are available upon reasonable request to the authors.

## (vi) Acknowledgments

The authors would like to thank “Forum Söder” for their help in retrieving data.

## Author contribution

Conception and design of study: SM, MP, MI, MS, AP, CC. Acquisition of data: SM, MP. Analysis and/or interpretation of data: SM, MP. Drafting of the manuscript: SM, MP. Revising the manuscript critically for important intellectual content: SM, MP, MI, MS, AP, CC. All authors approved the final version of the manuscript to be published.

## References

1. Zhu N, Zhang D, Wang W, Li X, Yang B, Song J, et al. A Novel Coronavirus from Patients with Pneumonia in China, 2019. N Engl J Med. 2020;382(8):727–33.

2. World Health Organization. WHO Coronavirus (COVID-19) Dashboard [Internet]. Geneva: World Health Organization; 2020 [cited 2021 Nov 7]. Available from: https://covid19.who.int.

3. European Centre for Disease Prevention and Control. Infection prevention and control and preparedness for COVID-19 in healthcare settings – Sixth update. Stockholm: ECDC; 2021 Feb.

4. World Health Organization. Recommendations for national SARS-CoV-2 testing strategies and diagnostic capacities. Geneva: World Health Organization; 2021 June.

5. Turcato G, Zaboli A, Pfeifer N, Sibilio S, Tezza G, Bonora A, et al. Rapid antigen test to identify COVID-19 infected patients with and without symptoms admitted to the Emergency Department. Am J Emerg Med. 2021;51:92–7.

6. Treggiari D, Piubelli C, Caldrer S, Mistretta M, Ragusa A, Orza P, et al. SARS-CoV-2 rapid antigen test in comparison to RT-PCR targeting different genes: A real-life evaluation among unselected patients in a regional hospital of Italy. J Med Virol. 2021.

7. Fournier PE, Zandotti C, Ninove L, Prudent E, Colson P, Gazin C, et al. Contribution of VitaPCR SARS-CoV-2 to the emergency diagnosis of COVID-19. J Clin Virol. 2020;133:104682.

8. Fitoussi F, Dupont R, Tonen-Wolyec S, Bélec L. Performances of the VitaPCR™ SARS-CoV-2 Assay during the second wave of the COVID-19 epidemic in France. J Med Virol. 2021;93(7):4351–7.

9. Region Skåne. Situation Report COVID-19 in Skåne. [Internet]. Region Skåne; 2020 [cited 2021 Nov 9]. Available from: https://www.skane.se/digitala-rapporter/lagesbild-COVID-19-i-skane/situation-report-in-english/#Cases.

10. World Health Organization. WHO COVID-19: Case defintions. Geneva: World Health Organization; 2020 Dec.

11. Bristol AA, Schneider CE, Lin SY, Brody AA. A Systematic Review of Clinical Outcomes Associated With Intrahospital Transitions. J Healthc Qual. 2020;42(4):175–87.

12. Webster J, New K, Fenn M, Batch M, Eastgate A, Webber S, et al. Effects of frequent PATient moves on patient outcomes in a large tertiary Hospital (the PATH study): a prospective cohort study. Aust Health Rev. 2016;40(3):324–9.

13. Blay N, Roche MA, Duffield C, Gallagher R. Intrahospital transfers and the impact on nursing workload. J Clin Nurs. 2017;26(23-24):4822–9.

14. Morley C, Unwin M, Peterson GM, Stankovich J, Kinsman L. Emergency department crowding: A systematic review of causes, consequences and solutions. PLoS One. 2018;13(8):e0203316.

